# SATB2/elastic lamina dual-staining in colon cancer: clinicopathological impact and prognostic value

**DOI:** 10.64898/2026.02.19.26346607

**Authors:** Bingjing Jiang, Yanfei Zhang, Huichao Sheng, Qinghua Wang, Bin Hu, Lixia Wang, Jianfei Fu

**Affiliations:** Affiliated Jinhua Hospital, Zhejiang University School of Medicine, jinhua, China, Department of Pathology; Affiliated Jinhua Hospital, Zhejiang University School of Medicine, jinhua, China, Department of Medical Oncology

**Keywords:** Colon cancer, Specific AT sequence binding protein 2, Elastic lamina, Dual-staining

## Abstract

**Objective:** To explore the application value of dual-staining for specific AT sequence binding protein 2 (SATB2) immunohistochemistry and elastic lamina in detecting elastic lamina invasion (ELI) in pT3 colon cancer, and to assess its association with clinicopathological characteristics, staging, and prognosis.

**Methods:** This retrospective cohort study enrolled 176 pT3 colon cancer patients who underwent radical resection at Affiliated Jinhua Hospital Zhejiang University School of Medicine. The deepest tumor-infiltrated paraffin blocks were collected for SATB2 immunohistochemistry and elastin dual-staining. Correlations between ELI status and clinicopathological characteristics and prognosis were analyzed. Survival data of 74 pT4a stage patients were collected for comparative analysis.

**Results:** ELI (+) was positively associated with high tumor budding grade, vascular invasion, lymph node metastasis, and reduced tumor infiltrating lymphocytes (TILs) (all *P* < 0.001). No correlations were observed with age, gender, tumor location, histological subtype, tumor grade, or perineural invasion (all *P* > 0.05). The ELI (+) group exhibited significantly shorter disease-free survival (DFS) and overall survival (OS) compared to ELI (-) group (*P* < 0.05). Additionally, the ELI (+) group demonstrated inferior OS than the pT4a group, though DFS did not differ significantly.

**Conclusion:** Dual-staining of SATB2 immunohistochemistry and elastic lamina provides a reproducible and objective method for assessing ELI. ELI correlates with key clinicopathological features and functions as an independent adverse prognostic indicator in pT3 colon cancer.

## Introduction

Colon cancer represents a predominant malignancy within the gastrointestinal tract. Recent data indicate that colon cancer is the third most commonly diagnosed malignancy worldwide and the second leading cause of cancer-related death^[1]^. While advances in precision medicine and multimodal therapeutic strategies have reduced mortality rates, the 5-year survival rate for advanced stages remains suboptimal, particularly in pT3 colon cancer patients where diverse clinicopathological high-risk factors contribute to heterogeneous outcomes. This prognostic uncertainty underscores the critical need for novel biomarkers to refine risk stratification. The TNM staging system, jointly defined by the American Joint Committee on Cancer (AJCC) and the Union for International Cancer Control (UICC), serves as the cornerstone for prognostic stratification and therapeutic decision-making in colon cancer^[2]^. Under the 8th edition of the AJCC colon cancer staging criteria, pT4a is defined as tumor invasion of the visceral peritoneum (VPI), including cases where the entire intestine passes through a tumor caused perforation, allowing the tumor to infiltrate the visceral peritoneal surface via an inflammatory pathway. Our previous study, utilizing public-access data, revealed that pT4a patients have a worse prognosis than pT3 patients and that the pT stage is closely linked to treatment selection^[3]^.

In practice, VPI assessment faces significant challenges^[4, 5]^. The fat-surrounded serosal membrane is hard to identify, and suspected serosal infiltration sites often involve the abdominal wall fold and extraperitoneal structures, making precise serosa localization in specimens difficult for pathologists^[6]^. Additionally, tumors near the serosa often induce structural damage, inflammation, mesothelial hyperplasia, and fibrosis. They also recruit macrophages (e.g., CD68 and CD204-positive) to the serosal vicinity, which can obscure normal serosal landmarks and lead to frequent inaccuracies in VPI identification^[6, 7]^. Consequently, the diagnostic accuracy and reproducibility of VPI have long been debated^[4–6, 8–11]^. Given the critical yet difficult nature of colon cancer pathological staging, identifying objective interpretation indicators for VPI is an urgent priority.

Given these challenges, elastic lamina has gained significant attention. In lung cancer staging, tumor invasion of the visceral pleural elastic lamina has been used as a criterion for pleural invasion since 1988 and has been widely adopted^[12, 13]^. The peritoneum and pleura share similar anatomical features. In colon cancer, elastic lamina invasion (ELI) is also an important pathological feature. Previous studies have demonstrated that ELI is associated with poor prognosis in colon cancer patients and is closely linked to tumor recurrence, metastasis, and prognosis. ELI can serve as a prognostic indicator for colon cancer^[6, 14–17]^. Some studies suggest that patients with ELI-positive pT3 and pT4a colon cancer should receive similar treatments^[18]^. ELI is expected to become a new anatomical marker replacing traditional visceral pleural invasion diagnosis.^[4, 14, 15, 17, 19–21]^。

However, existing studies often have small sample sizes and commonly use single elastic lamina staining. Inflammatory cell infiltration and fibrous tissue hyperplasia often make tumor tissues, especially scattered tumor cells, nearly indistinct. Consequently, the relationship between tumors and elastic lamina is difficult to clarify, which affects the accuracy of ELI results. Their presence is frequently underestimated in pathological evaluations, impacting the accurate assessment of tumor invasiveness and prognosis. Hence, developing an effective and accurate detection method is crucial for enhancing the diagnosis and treatment of colon cancer. To address this, we have explored a dual-staining method combining immunohistochemistry and elastic lamina staining.

## 1 Materials and Methods

### 1.1 Study design, ethical approval and consent

This is a retrospective cohort study. The recruitment period for patient identification and data collection spanned from January 2017 to December 2022. The study protocol was reviewed and approved by the Medical Ethics Review Committee of Jinhua Central Hospital (Approval No. (Research) 2022-Ethics Review-33). The requirement for written informed consent was waived by the ethics committee due to the retrospective nature of the study, which involved the analysis of anonymized archival pathological specimens and clinical data.

### 1.2 Study cohort and data acquisition

Using the pathology workstation (EW PIS V5.0) of Affiliated Jinhua Hospital, Zhejiang University School of Medicine, we retrieved cases of patients who underwent radical resection for colon cancer from January 2017 to December 2022. We selected 176 pT3 stage II-III colon cancer patients who did not receive neoadjuvant therapy. Their clinical characteristics, postoperative treatment, and prognosis data were obtained via the Haitai Electronic Medical Record System (V3.0) and the hospital’s colon cancer database (GPS5.0). Additionally, 74 pT4a stage II-III colon cancer patients who did not receive neoadjuvant therapy were selected to form the pT4a group. Data on adjuvant treatment receipt and survival outcomes were collected.

### 1.3 Histopathological review and case selection

All hematoxylin-eosin (HE)-stained sections underwent blinded independent review by two gastrointestinal-specialized pathologists. Under a double-blind protocol, reviewers assessed sections while blinded to clinical outcomes and recorded the results. Discrepancies were resolved through multi-headed microscopy consensus. Cases were included if they met all the following criteria:

- Histologically confirmed invasive colon adenocarcinoma.
- Good quality samples with intact layers.
- Deepest invasion reaching the subserosal layer (pT3).

The deepest tumor-invaded paraffin blocks were selected and Tumor-Infiltrating Lymphocytes (TILs) values were quantified at the invasive margin following the International TILs Working Group (ITWG) consensus guidelines^[22]^. Specifically, TILs were defined as the percentage of mononuclear cells (lymphocytes and plasma cells) in the stromal area at the tumor invasion front on HE-stained colon cancer sections. TILs ≤ 20% indicated low TILs, while TILs > 20% indicated high TILs^[23]^.

### 1.4 Dual-staining for specific AT sequence binding protein 2 (SATB2) immunohistochemistry and elastic lamina

#### 1.4.1 Dewaxing and hydration

Sections were dewaxed in fresh xylene for 15 minutes twice, then transferred to absolute ethanol for 3 minutes twice, followed by 95% ethanol for 3 minutes, 85% ethanol for 3 minutes, and rinsed with tap water for 1 minute. Finally, they were washed with PBS 3 times, 3 minutes each.

#### 1.4.2 Antigen retrieval with EDTA

Sections were placed on a high-temperature staining rack and put into a boiling EDTA antigen retrieval solution in a stainless-steel pot. The sections were heated for 20 minutes under minimal power, then removed from the heat and cooled naturally for 10 minutes. Tap water was used to cool the pot’s exterior. Once the solution reached room temperature, the sections were removed, rinsed with distilled water 3 times for 3 minutes each, and washed with PBS three times for 3 minutes each.

#### 1.4.3 Blocking endogenous peroxidase

Remove the PBS, add 100 μl of endogenous peroxidase blocker, and incubate at room temperature for 10 minutes. Rinse with PBS 3 times, 30 seconds each.

#### 1.4.4 Adding primary antibody

Remove the PBS, add 100 μl of primary antibody, and incubate at room temperature for 60 minutes. Rinse with PBS 3 times, 30 seconds each.

#### 1.4.5 Adding enzyme-labeled polymer

Remove the PBS, add 100 μl of enzyme-labeled goat anti-mouse/rabbit IgG polymer, and incubate at room temperature for 15 minutes. Rinse with PBS 3 times, 30 seconds each.

#### 1.4.6 Diaminobenzidine (DAB) staining

Remove the PBS, add 100-200 μl of freshly prepared DAB solution, and incubate for 3-5 minutes. Observe the staining result under a light microscope, usually within 10 minutes.

#### 1.4.7 Elastic lamina staining

Rinse with 95% alcohol. Place the sections in elastin staining solution at room temperature (8-24 hours) with the container covered. Quickly rinse with 95% alcohol to remove excess stain, then wash with water. Perform alcohol dehydration, xylene clearing, and mount with neutral gum.

### 1.5 Microscopic diagnosis

Two gastrointestinal-specialized pathologists with extensive diagnostic experience performed a double-blind assessment of the slides under a microscope and recorded the results. Discrepant cases were discussed among multiple pathologists to reach a consensus. The ELI results were documented and statistically analyzed.

### 1.6 Grouping and statistical analysis

Statistical analysis was conducted using software such as R and Excel. Specimens demonstrating tumor penetration of the elastic lamina were classified as ELI (+), whereas those without penetration were classified as ELI (−). Cases lacking an identifiable elastic lamina were designated EL (−). For subsequent analyses, ELI (+) samples constituted the ELI (+) group, while ELI (−) and EL (−) cases were combined into a single ELI (−) group^[24]^. Survival data for both groups were retrieved from our hospital’s GPS database. Survival data of 74 pT4a stage patients served as the control group. Kaplan-Meier curves were plotted, and cox proportional hazards regression was used for analysis. Clinical and pathological characteristics (age, gender, tumor location, histological subtype, tumor grade, tumor budding grade, vascular invasion, perineural invasion, and N stage) of the ELI (+) and ELI (-) groups were extracted from the database. These data were verified under the microscope and combined with TILs values for statistical analysis. A comparative analysis was performed on the receipt of postoperative treatment between the ELI (+) group and pT4a group.

## 2 Results

### 2.1 Cohort characteristics and morphological assessment

A cohort of 176 pT3 colon cancer cases was initially enrolled. Following exclusion of 3 cases (1.7%) due to specimen processing failure (tissue fragmentation), 173 cases underwent successful dual-staining analysis. Final stratification revealed: 93 patients ELI (–), 5 patients EL (–) and 75 patients ELI (+) (details in Fig 1). Specimens were reclassified into two groups: ELI (–) group (n=98 patients) and ELI (+) group (n=75 patients), the former comprising all ELI (–) cases (n=93 patients) together with EL (–) cases (n=5 patients). The microscopic morphologies were shown in Figs 2–4.

**Figure 1.**
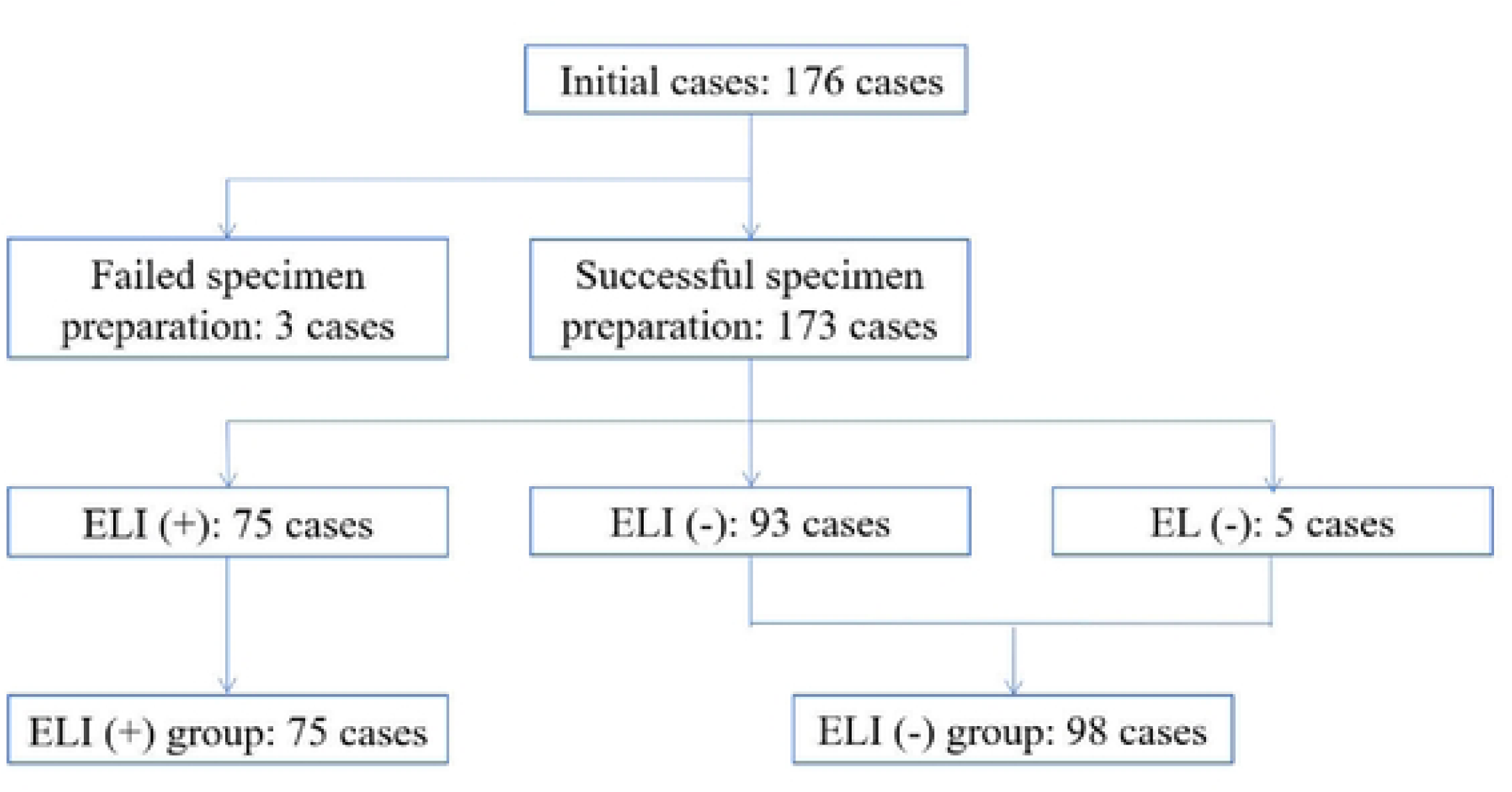
Flowchart of case selection and grouping in the study.

**Figure 2.**
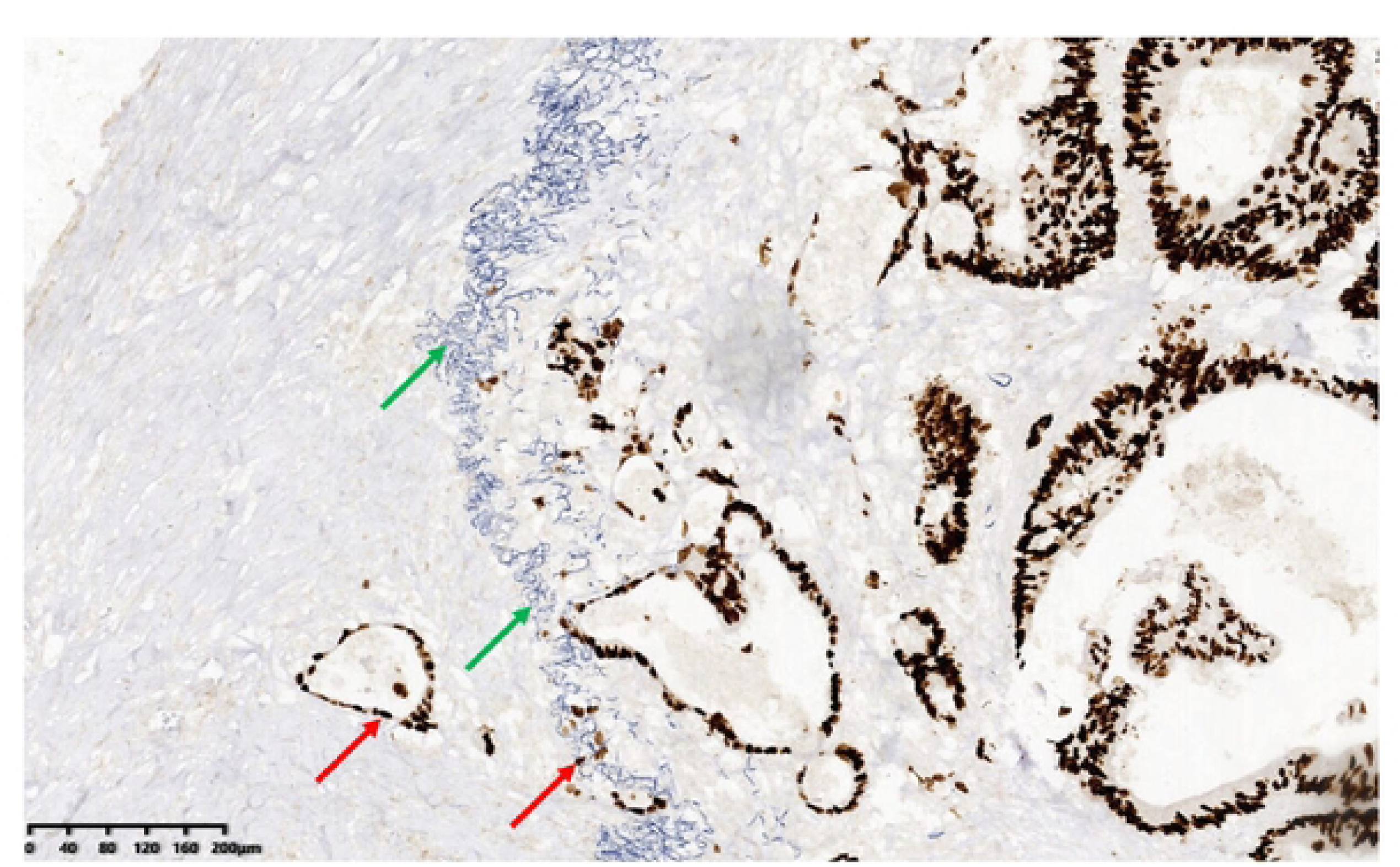
Dual-staining section of the ELI(+) group under microscopy (×100) Brown tissue (red arrows) indicates SATB2 immunohistochemistry-positive colon adenocarcinoma cell nuclei. Blue linear tissue (green arrows) denotes the elastic lamina. Tumor cells infiltrate from right to left, invade and penetrate the elastic lamina. Scattered individual tumor cells breaching the elastic lamina can be clearly identified.

**Figure 3.**
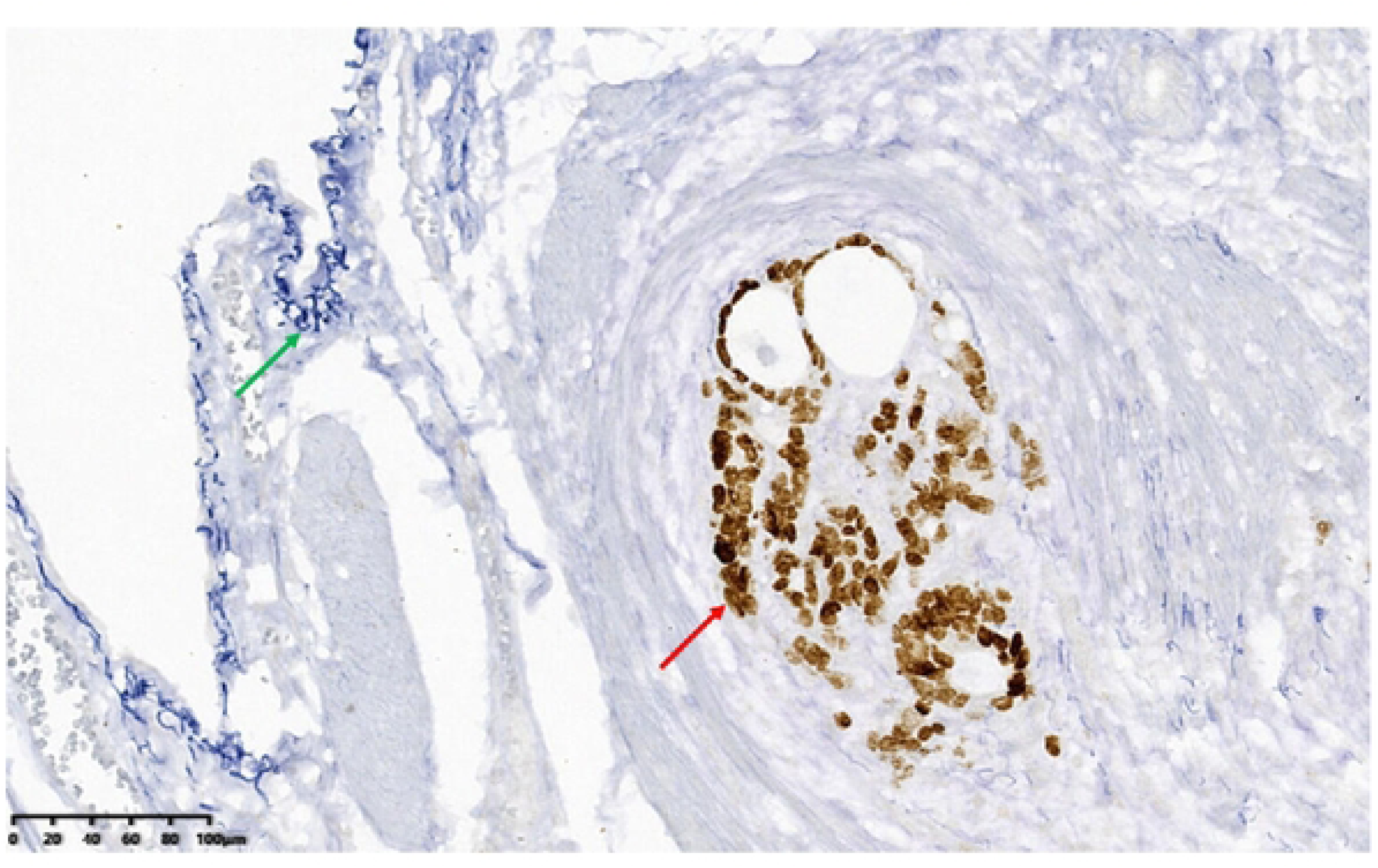
Dual-staining section of the ELI(-) group under microscopy (×200). Brown tissue (red arrow) marks SATB2 immunohistochemistry-positive colon adenocarcinoma cell nuclei. Blue linear tissue (green arrow) denotes the elastic lamina. Tumor cells infiltrate upward but don’t breach the elastic lamina.

**Figure 4.**
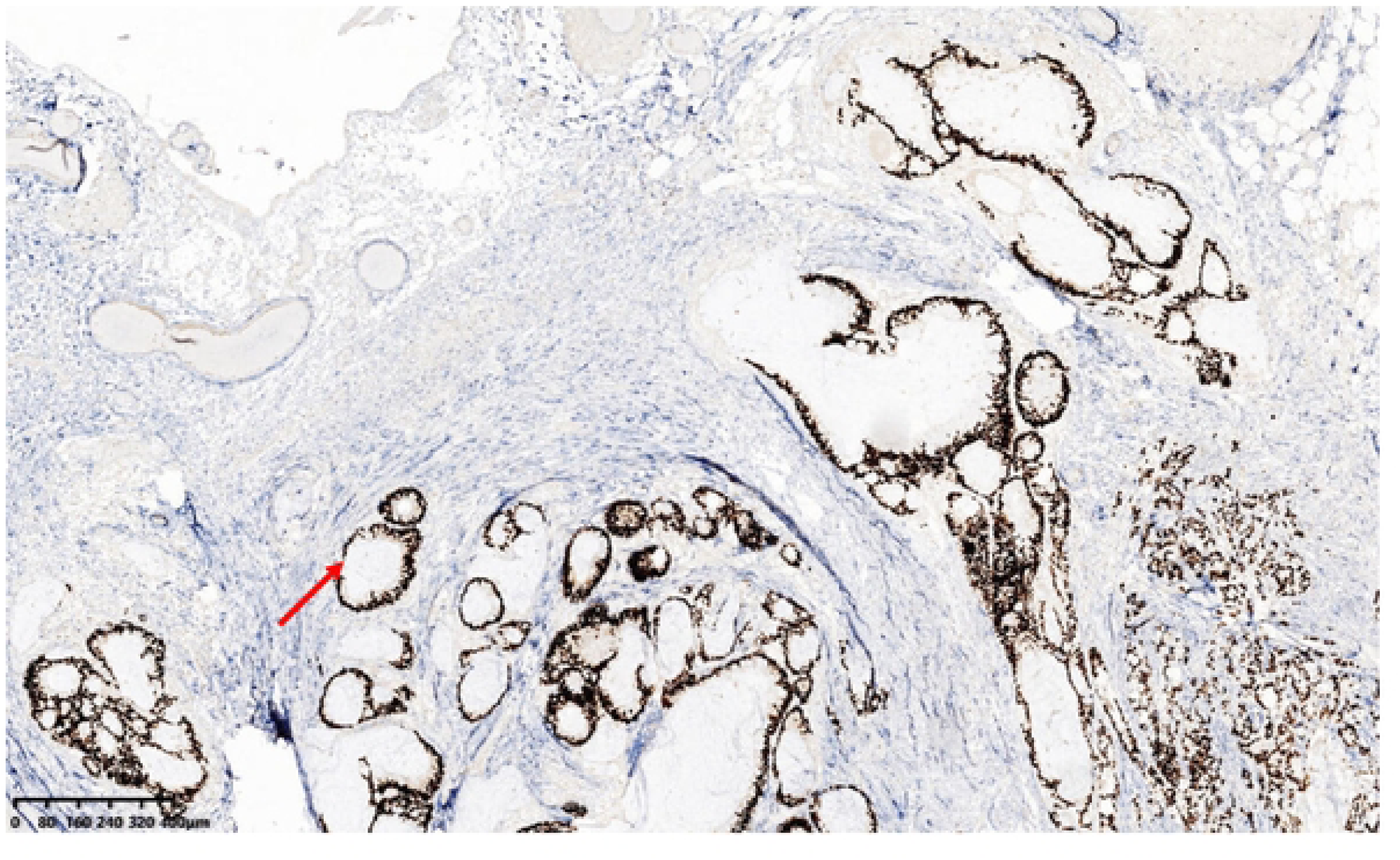
Dual-staining section of the EL(-) group under microscopy (×40). Brown tissue (red arrow) marks SATB2 immunohistochemistry-positive colon adenocarcinoma cell nuclei with absent elastic lamina.

### 2.2 Association between ELI status and clinicopathological characteristics of colon cancer

ELI (+) showed a positive correlation with high tumor budding grade, vascular invasion, lymph node metastasis, and reduced TILs (all *P* < 0.001). However, no significant statistical correlation was found between ELI (+) and age, gender, tumor location, histological subtype, tumor grade, or perineural invasion (all *P* > 0.05). Comprehensive clinicopathological characteristics stratified by ELI status are presented in Table 1.

**Table 1.**
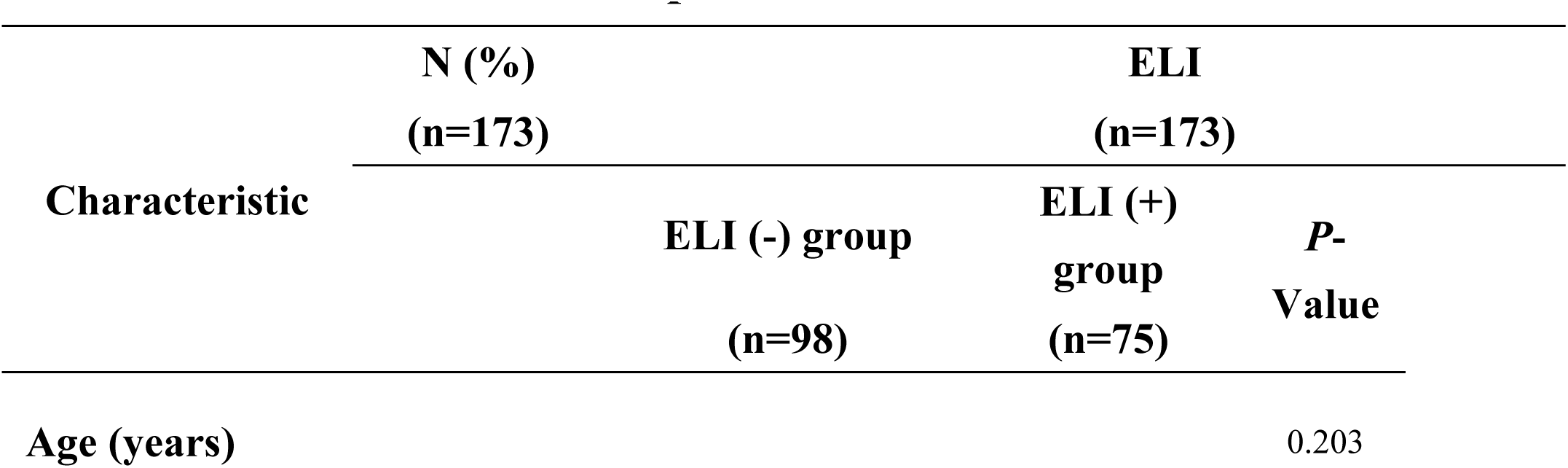
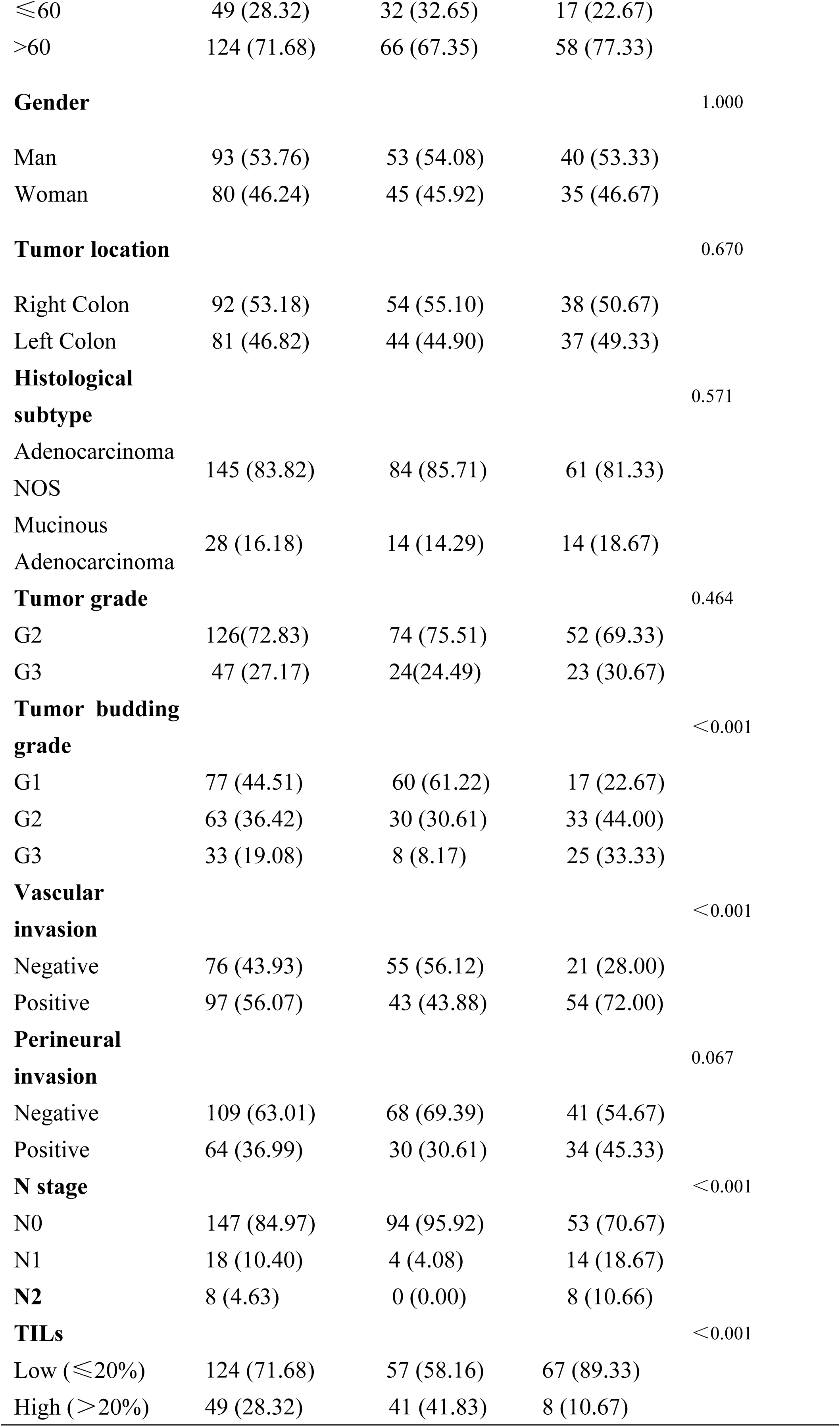
Clinicopathological characteristics and ELI status of the 173 colon.

### 2.3 Correlation between ELI status with the stage and prognosis of colon cancer

#### 2.3.1 Comparative survival analysis: ELI (+) group vs ELI (-) group

Kaplan-Meier analysis with log-rank testing demonstrated significantly inferior survival outcomes in ELI (+) group patients compared to the ELI (−) group patients. Disease-free survival (DFS) was significantly shorter in ELI (+) group patients (HR = 2.18, 95% CI: 0.99–4.77, *P* = 0.009; Fig 5A). Similarly, overall survival (OS) was significantly reduced in the ELI (+) group (HR = 3.35, 95% CI: 1.10–10.15, *P* = 0.029; Fig 5B).

**Figure 5A.**
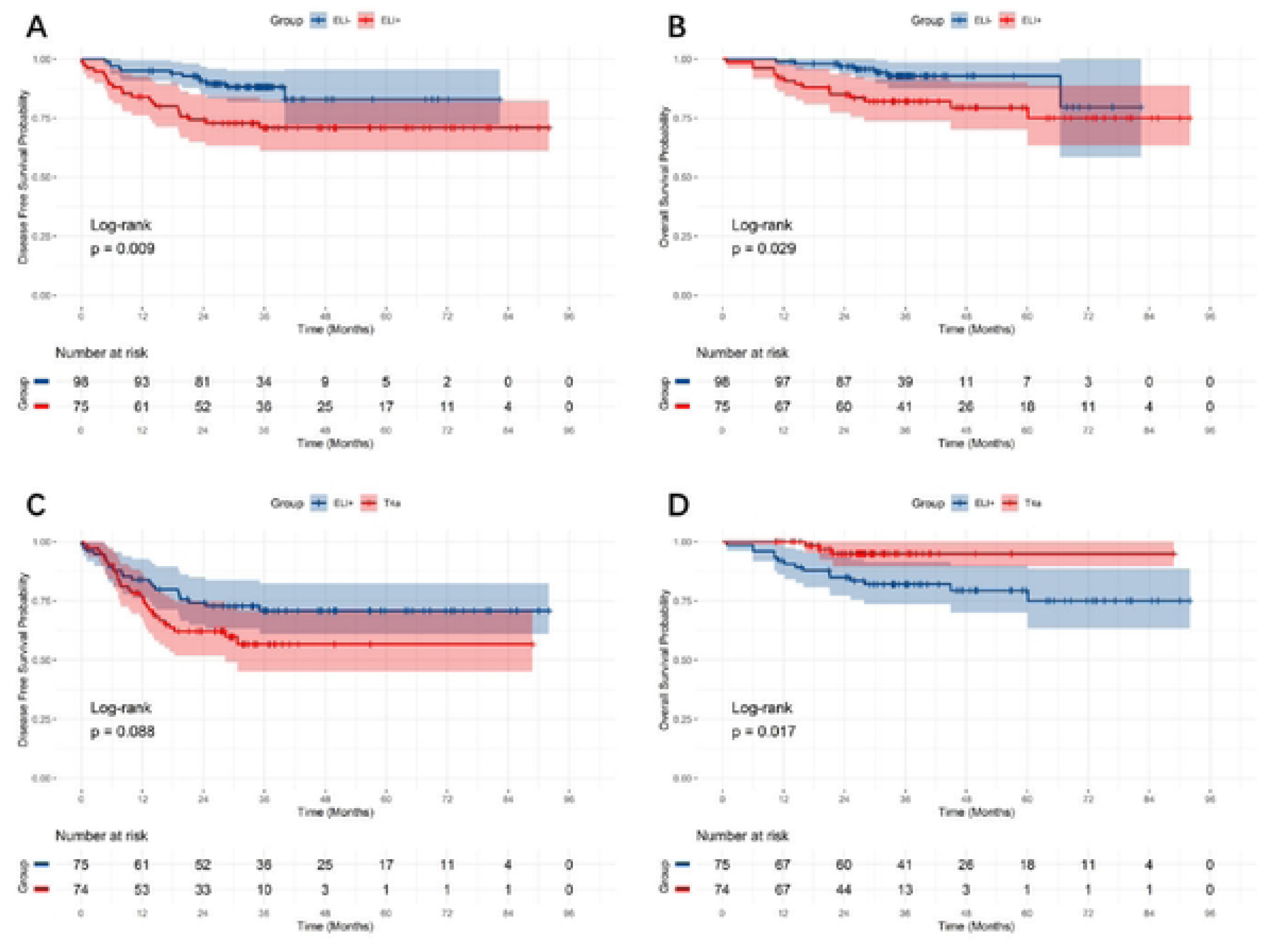
5B Kaplan-Meier curves ofDFS and OS for the ELI(-) and ELI(+) groups. Figure 5C, 5D Kaplan-Meier curves ofDFS and OS for the ELI(+) and pT4a groups.

#### 2.3.2 Comparative survival analysis: ELI (+) group vs pT4a group

DFS was marginally longer in the ELI (+) group than in the pT4a group, but the difference did not reach statistical significance (HR = 0.61, 95% CI: 0.35–1.08, *P* = 0.088; Fig 5C). Conversely, OS was significantly superior in the pT4a group (HR = 0.24, 95% CI: 0.07–0.85, *P* = 0.017; Fig 5D).

#### 2.3.3 Tripartite survival stratification: ELI (-) group vs ELI (+) group vs pT4a group

Survival analysis demonstrated a significant hierarchical stratification among the groups. For DFS, the ELI (−) group exhibited the most favorable outcome, followed by the ELI (+) and pT4a groups (*P* < 0.0001; Fig 6A). A similar gradient was observed for OS, with the ELI (−) group showing the best prognosis, followed by the pT4a group, while the ELI (+) group demonstrated the least favorable outcome (*P* = 0.022; Fig 6B).

**Figure 6A.**
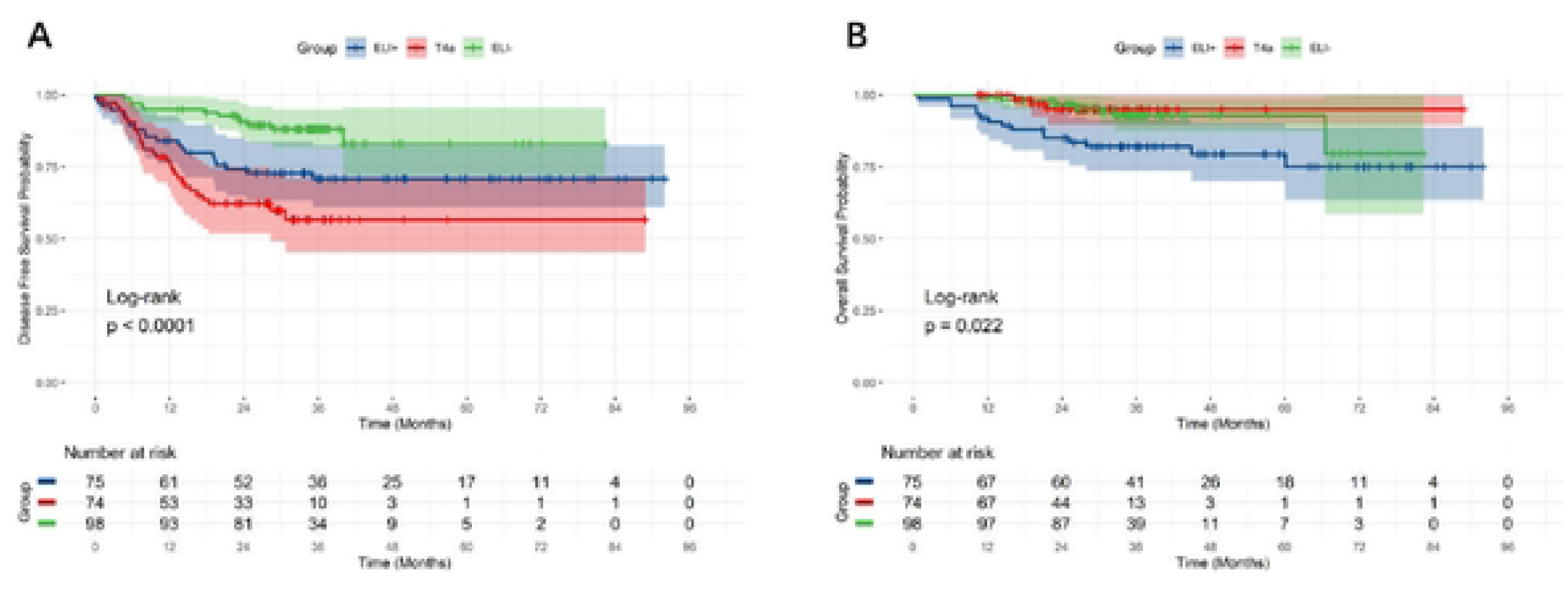
Kaplan-Meier curve ofDFS for the ELI(-), ELI(+), and pT4a groups. Figure 6B Kaplan-Meier curve of OS for the ELI (-), ELI (+), and pT4a groups.

### 2.4 Disparities in the receipt of postoperative treatment: ELI (+) group vs pT4a group

A significantly lower proportion of patients in the ELI (+) group received postoperative treatment compared to those in the pT4a group (*P* < 0.001; Table 2).

**Table 2.**
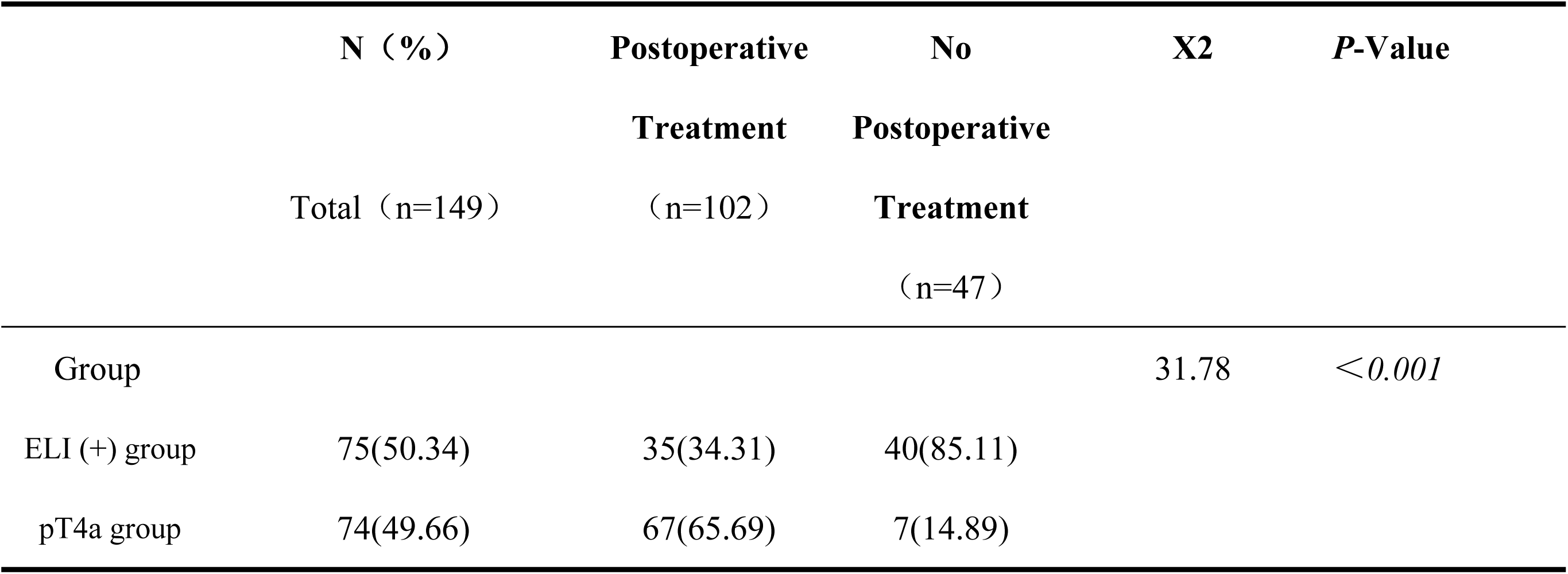
Disparities in postoperative treatment proportion: ELI (+) group vs pT4a group.

## 3 Discussion

Colon cancer remains a major cause of cancer-related mortality worldwide. Precise evaluation of tumor invasiveness and prognosis is crucial for guiding postoperative treatment strategies and represents a critical focus of ongoing research. Identifying high-risk patients who are most likely to benefit from personalized therapeutic interventions is essential for improving clinical outcomes.

SATB2, a nuclear matrix protein, serves as a highly specific and sensitive immunohistochemical marker for colon cancer tumor cells^[25, 26]^. In this study, we innovatively integrated SATB2 immunohistochemistry with elastic fiber staining to develop a dual-staining protocol. This method simultaneously visualizes tumor cells and the elastic lamina architecture, enabling precise and reliable identification of ELI. Compared to traditional single-stain elastic lamina staining, it better distinguishes tumor cells from the surrounding fibrous matrix, avoiding misjudgment from inflammation or fibrosis. This improvement provides a novel tool for colon cancer staging and treatment, and promotes standardized ELI evaluation.

Notably, 5 (2.9 %) specimens lacked an identifiable elastic lamina, designated as EL (−), in line with earlier observations^[21, 24]^. This phenomenon may be attributed to several factors. Anatomically, the peritoneal elastic lamina is not circumferentially continuous along the intestinal wall and is often discontinuous or terminates adjacent to the posterior abdominal wall, resulting in segmental absence^[27]^. Additionally, natural interindividual variation can lead to regions where the elastic lamina is microscopically thin, fragmented, or entirely missing. Technical aspects such as mechanical injury during specimen retrieval or suboptimal processing steps (e.g., inadequate fixation, dehydration or embedding) may also compromise elastic lamina integrity. Accordingly, current guidelines recommend examining multiple tissue blocks for elastic lamina evaluation and classifying cases with demonstrable absence or discontinuity as ELI (−)^[21, 24]^. This study adhered to these established criteria.

ELI (+) was significantly associated with adverse clinicopathological features, including high tumor budding grade, vascular invasion, lymph node metastasis, and reduced TILs (all *P* < 0.001), suggesting that ELI (+) lesions harbor an aggressive phenotype and unfavorable biology, in partial agreement with Kojima et al.^[6]^. Histologically, ELI (+) areas are characterized by dense fibrosis and a distinct tumor microenvironment enriched in chemokines, cytokines, and growth factors that can trigger epithelial–mesenchymal transition (EMT) and suppress local immune responses^[6, 28]^. Tumor budding, a surrogate marker of EMT, reflects enhanced metastatic potential, whereas diminished TILs indicate an immunosuppressive milieu. Their combined effect likely underpins the poor prognosis observed in ELI (+) patients and provides novel perspectives for investigating the immune landscape of colon cancer. These findings align with recent reports identifying ELI as an independent adverse prognostic factor, underscoring its utility for objective risk stratification of pT3 colon cancer patients^[24]^.

In a seminal study, Bi et al. examined 105 colon cancer specimens and demonstrated that pT3 patients with ELI (+) exhibited significantly shorter DFS than those with ELI (−), and that ELI (+) independently predicted recurrence after surgery for stage III disease^[20]^. Our present cohort corroborates these findings: both DFS and OS were significantly reduced in pT3 ELI (+) patients compared with ELI (−) controls, confirming ELI (+) as a robust indicator of adverse outcome. Based on these results, we propose incorporating ELI into the subclassification of pT3 tumors, distinguishing pT3/ELI (−) from pT3/ELI (+) to better identify high-risk subsets within this stage. Prior investigations have reported that outcomes for pT3 ELI (+) stage II tumors resemble those of pT4a lesions, suggesting that pT3/ELI (+) may represent a biological entity similar to pT4a disease^[14, 15, 24]^. Although DFS in the ELI (+) group was marginally higher than that in the pT4a group, the difference did not reach statistical significance (*P* = 0.088). In contrast, OS was significantly shorter in the pT3 ELI (+) group than in the pT4a group. This discrepancy may be attributed to differences in postoperative treatment proportion: only 46.7 % of pT3 ELI (+) patients received postoperative treatment compared with 90.5 % of pT4a patients, who-by virtue of their higher stage-were treated more aggressively, likely mitigating otherwise adverse outcomes. Larger prospective studies with longer follow-up are warranted to validate this hypothesis. Collectively, these findings underscore that ELI serves not only as a robust pathological prognosticator but also as a potential biomarker to guide postoperative treatment decisions in pT3 colon cancer.

Integrating SATB2 immunohistochemistry with elastic lamina dual-staining refines assessment of invasion depth, recurrence risk, and patient outcome, enabling truly individualized treatment planning.

Several limitations should be considered in this study, including its modest sample size and retrospective single-center design, which may limit the generalizability of the findings. Future multi-center studies with larger patient cohorts, augmented by complementary molecular markers and advanced imaging, are warranted to validate ELI-based risk stratification. Moreover, application of single-cell sequencing or spatial transcriptomics to ELI (+) regions could help elucidate the molecular determinants linking ELI to metastasis and immune evasion, further refining precision oncology for colon cancer.

## 4 Conclusion

The dual-staining technique for SATB2 immunohistochemistry and elastic lamina offers a reproducible and objective approach for evaluating ELI. ELI correlates with key clinicopathological features and serves as an independent adverse prognostic indicator in pT3 colon cancer, underscoring its potential value in guiding postoperative treatment strategies. However, these findings are limited by the modest sample size and single-center design. Thus, future prospective multi-center studies with larger cohorts are warranted. Integration of complementary molecular markers, high-resolution imaging, and emerging technologies such as single-cell sequencing or spatial transcriptomics will further refine ELI detection, clarify its mechanistic links to metastasis and immune evasion, and promote precision management of colon cancer.

## Data Availability

All relevant data are within the manuscript and its Supporting Information files.

## Declarations

## Ethics approval and consent to participate

This study was performed in line with the principles of the Declaration of Helsinki. Approval was granted by the Medical Ethics Review Committee of Jinhua Central Hospital (Approval No. (Research) 2022-Ethics Review-33). The need for written informed consent was waived by the Medical Ethics Review Committee of Jinhua Central Hospital due to the retrospective nature of the study.

## Consent for publication

Not applicable.

## Availability of data and materials

The datasets used and analyzed during the current study are available from the corresponding author on reasonable request.

## Competing interests

The authors declare that they have no competing interests.

## Funding

The study was supported by the The General Program of Jinhua Municipal Science & Technology Bureau (grant number: 2023-4-070) and The Key Program of the Affiliated Jinhua Hospital, Zhejiang University School of Medicine (grant number: JY2021-1-05).

## Authors’ contributions

BJ: Methodology, Investigation, Validation, Writing– Original Draft Preparation, Funding Acquisition, Project Administration. YZ.HS.BH: Pathological Techniques, Resources. QW: Formal Analysis, Data Curation, Visualization. LW: Investigation, Resources, Supervision. JF: Conceptualization, Methodology, Formal Analysis, Writing – Review & Editing.

## Acknowledgements

Not applicable.

## Notes

### Competing Interest Statement

The authors have declared no competing interest.

### Funding Statement

Yes

### Author Declarations

This study was performed in line with the principles of the Declaration of Helsinki. Approval was granted by the Ethics Committee of Affiliated Jinhua Hospital, Zhejiang University School of Medicine (Ethical approval number: 2022-33). The need for written informed consent was waived by the Ethics Committee of Affiliated Jinhua Hospital, Zhejiang University School of Medicine due to the retrospective nature of the study.

